# Personalized volume-deescalated elective nodal irradiation in oropharyngeal squamous cell carcinoma (DeEscO): a study protocol

**DOI:** 10.1101/2025.06.24.25330129

**Authors:** Esmée L Looman, Yoel Pérez Haas, Roman Ludwig, Debra Fesslmeier, Grégoire B Morand, Matthias Guckenberger, Olgun Elicin, Roland Giger, Sébastien Tran, Francesco Martucci, Sarah Melab-Belkhodja, Oliver Riesterer, Stephan Benke-Bruderer, Jan Unkelbach, Panagiotis Balermpas

## Abstract

**Background:** Definitive (chemo)radiotherapy of oropharyngeal squamous cell carcinoma (SCC) consists of treatment of the macroscopic tumor and prophylactic irradiation of large parts of the lymphatic drainage system: the elective clinical target volume (CTV). There is limited data and quantification of (occult) lymphatic spread and the required extent of the elective CTV. De-escalation of the irradiated volume could result in less early and late toxicity and less affection of the immune system. In this study, we reduce the elective CTV based on the personalized factors T-stage, clinical lymphatic involvement, and lateralization of the primary tumor.

**Methods:** The primary objective is to evaluate the efficacy and safety of an individualized de-escalation of elective node irradiation volumes. We collected a multi-institutional dataset of 598 oropharyngeal SCC patients in whom detailed lymph node involvement was reported. The data can be explored on the publicly available online platform LyProX.org. Based on the data, we developed a model of lymphatic tumor progression to estimate the probability of occult metastases in the clinically negative lymph node levels (LNLs). Metastatic lymphatic progression is described via a hidden Markov model. The patient’s state of lymph node involvement is described by hidden binary random variables, and the transition matrix describes the probabilities of lymphatic spread. Based on this model and clinical judgement, we created tables that define the personalized elective CTV for different combinations of T-stage, midline extension and clinical LNL involvement. The elective CTV is defined such that the cumulative risk of occult metastases in all non-irradiated LNLs is below 10%. The primary endpoint of this multicentric, prospective, single-arm trial is the rate of out-of-field nodal recurrences after 2 years. The study will be conducted in 6 centers across Switzerland and 120 patients will be enrolled.

**Discussion:** This study sets out to personalize the elective CTV in oropharyngeal SCC patients, with the goal to de-escalate radiotherapy for patients and herewith improve therapy-associated toxicity and quality of life. The results of this study may be used to personalize treatment in oropharyngeal SCC patients.

**Trial Registration:** This study sets out to personalize the elective CTV in oropharyngeal SCC patients, with the goal to de-escalate radiotherapy for patients and herewith improve therapy-associated toxicity and quality of life. The results of this study may be used to personalize treatment in oropharyngeal SCC patients.

## Background

Curative intended treatment of squamous cell carcinoma (SCC) of the oropharynx can consist of surgery, (chemo)radiotherapy, or a combination of both. When treated with radiation, the target volume contains not only the primary tumor and clinically detected lymph node metastases. In addition, a large part of the lymphatic drainage system of the neck which is at risk of harboring occult metastases, is irradiated - the so called “elective clinical target volume (CTV)” [1–3]. The current guidelines for the selection of lymph node target volumes are based on the AJCC/UICC 7^th^ edition classification of the nodal category regardless of the p16-status [3–5]. For p16+ oropharyngeal cancers, the guidelines mention to take the total number of positive lymph nodes, their size and their localization (ipsilateral, contralateral, and bilateral) into account for defining the low-risk nodal target volume selection. For p16 negative tumors, the levels to be included are determined by the nodal category [3]. The elective CTV is currently based on clinical recommendations, but there is only limited quantitative data available on (occult) lymphatic spread and the required size of the elective CTV [3], especially with regard to further personalization. This elective irradiation of the neck lymph node levels (LNL), however, is associated with early and late toxicity. Toxicities such as pain, dermatitis, mucositis, but also long-term sequela like swallowing dysfunction, xerostomia, lymphedema, thyroid dysfunction, and dysgeusia are commonly described, which potentially lead to hospitalization or long-term symptoms with subsequent life-quality impairment [6–11].

In addition, there is increasing interest in possible immune-sparing effects of radiotherapy de-escalation. Irradiation of the lymph nodes and lymph drainage system is associated with the development of lymphopenia [12]. The occurrence of (treatment-induced) lymphopenia and a possible impairment of the priming of T-cells are associated with a worse prognosis, possibly because of impairment of adaptive immune-mediated tumor responses [13, 14]. By volume de-escalation, the lymphatic system could be spared more, potentially leading to improved antigen-priming, less lymphopenia and better survival outcomes [15–17].

For standard-of-care (chemo)radiotherapy treatment in oropharyngeal SCC, treatment failure and out-of-field lymph node metastases have a low incidence. Leeman et al. showed a local treatment failure at 5 years of 4.2%, with 2.9% regional nodal failure and no marginal or isolated out-of-radiation-field failure [18]. Similar results were found by Van den Bosch et al., who found a 2-year recurrence rate in electively irradiated lymph node regions of 5.2% [19]. The low incidence of out-of-field lymph node recurrences suggests that there is potential for safely reducing the elective CTV without substantially increasing the risk of regional failure but reducing early and late toxicity.

We developed a statistical model to estimate the patients’ individual risk of occult lymph node metastases in clinically negative LNLs in newly diagnosed oropharyngeal SCC patients. This individual estimation is based on the patient’s distribution of macroscopic metastases, T-stage, and lateralization of the primary tumor [20, 21]. Based on tumor characteristics (T-stage, upstream nodal involvement, midline extension) and clinical experience, we personalize the elective clinical target volume based on a patient’s individualized risk. In this study, the elective CTV is defined such that the overall risk of occult lymph node metastases in all non-irradiated LNLs (out-of-field) is estimated to be below 10%.

The aim of this phase II clinical trial is to determine the efficacy and safety of the use of a personalized volume de-escalated elective nodal CTV in oropharyngeal SCC patients treated with primary (chemo)radiotherapy.

### Previous work on the development of a statistical model of occult lymph node involvement

We collected a multi-institutional dataset of 598 oropharyngeal SCC patients in whom the detailed patterns of lymph node involvement are reported. For each patient, lymph node involvement was recorded for each individual lymph node level (LNL) based on the available diagnostic modality. Pathology after neck dissection was reported for the subset of surgically treated patients. For most patients, clinical lymph node involvement based on imaging and fine needle aspiration (FNA) is reported. In addition, primary tumor location, lateralization, and T-stage is reported among other patient and tumor characteristics. The dataset is publicly available and is composed of data from three projects. A dataset of 287 patients from the University Hospital Zurich (USZ) are published and described in [22, 23]. The project was approved under BASEC-No. 2019–00684. A dataset of 74 oropharyngeal SCC patients from University Hospital Bern (ISB) is also published [24]. This project was approved under BASEC No. 2018–01716. The dataset of the remaining 237 oropharyngeal SCC patients from Centre Léon Bérard (CLB) in Lyon, France is described in the respective publication [23, 24]. In the original projects, only data from patients who signed a general consent and agreed to the further use of their data for research purposes was used, or for which approval under HRA art. 34 (without consent) was obtained. The publicly available online platform LyProX.org was developed to visualize published datasets in an intuitive and interactive manner. The tool including the publicly available data was used for the statistical analysis described below.

To define the elective nodal CTV, we are interested in the conditional probability that a clinically negative LNL harbors occult metastases, given the primary tumor characteristics and the location of clinically detected lymph node metastases. This conditional probability of occult disease depends on 1) the probability of the tumor to spread to the LNL of interest, and 2) the sensitivity and specificity of clinical diagnostic methods to detect lymph node metastases. To combine both aspects, we developed a comprehensive statistical model to predict the probability of occult disease per LNL using a Hidden Markov model (HMM). In this model, the state of involvement of a LNL is described by a latent binary random variable, which indicates if the LNL is healthy or harbors metastases (including occult metastases). The state of lymph node involvement of a patient is then described by a vector of binary random variables. Each LNL is further associated with an observed state, another binary random variable that indicates if a LNL is clinically diagnosed as involved. The HMM is used to describe the spread from the primary tumor to the LNLs and progression from an involved LNL to a downstream LNL. The model is parameterized using a directed graph where the arcs represent the possible spread to and between LNLs, resembling the anatomically defined lymph drainage of the neck. The main model parameters are the probabilities of spread associated with each arc. The latent true state of involvement and the clinically observed involvement are linked via sensitivity and specificity of imaging. Thus, we can compute the likelihood of the observed dataset of 598 patients given the model parameters. We use Markov Chain Monte Carlo (MCMC) sampling to draw these parameters from the posterior distribution, comprised of the likelihood and a uniform prior. This approach naturally provides uncertainty bounds for the model’s parameters and consequently also for the predicted risk of occult involvement. Model training is performed based on a consensus decision on the most likely state of involvement given all diagnostic modalities available for each patient and LNL in the dataset, which is assumed to represent the true latent state of involvement. Sensitivities and specificities are not learned, but taken from literature values, originating from the comparison of clinical and pathological involvement of patients receiving neck dissection. For risk calculations we set specificity to 1 and sensitivity to 0.81. This corresponds to the literature values for sensitivity of CT and MRI reported by De Bondt et al. [25]. A specificity of 1 corresponds to the assumption that clinically involved LNLs truly harbor metastases and do not represent false positives. A detailed description of the methodology can be found in the respective publications [20, 21, 26, 27]. The model presented in [26] is representative for the model underlying the design of the DeEscO trial, despite not being identical in all details.

The trained model can then be used for predicting the probability of occult involvement per LNL for individual patients. The patient characteristics that are input to the model are 1) clinical involvement per LNL (considering LNLs I, II, III, IV, V, and VII, both ipsilateral and contralateral), 2) early (T1/T2) versus advanced (T3/T4) T-stage, and 3) lateralization of the primary tumor (described as a binary variable that indicates if any part of the primary tumor extends beyond the midline). In addition, we considered central tumors, where there is no clear origin of the primary tumor lateral to the midline. For the common states of the primary tumor and clinical lymph node involvement (the patients that occurred at least once in the dataset), the model was used to predict the risk of occult disease in the clinically negative lymph node levels. To define the elective nodal CTV (CTV-3), the clinically negative LNLs are ranked according to their risk of occult involvement. The LNLs with the highest risk are included in the CTV-3 until the cumulative risk of involvement in all remaining LNLs is below 10% for 95% of the model parameter samples generated through MCMC. As a final measure of quality assurance, the CTV-3s constructed in this way have been discussed individually by the investigators, to ensure consistency with data and clinical judgement and experience.

### Objectives and Outcomes

The primary objective is to evaluate the efficacy and safety of personalized reduction of the elective nodal CTV in oropharyngeal SCC patients, based on the individual patient’s state of disease progression and risk factors, as measured by N-site recurrences in non-irradiated LNLs 2 years after finishing treatment.

Our hypothesis is that lymph node metastases in non-irradiated LNLs (out-of-field regional control) after personalized radiation treatment volume reduction occur in <10% of the participants.

Secondary objectives and the corresponding outcomes are to assess:

- Efficacy and safety of personalized reduction of the elective nodal CTV in oropharyngeal SCC patients, based on the individual patient’s state of disease progression and risk factors, as measured by N-site recurrences in non-irradiated LNLs 3 years after finishing treatment. This outcome will be measured by the Kaplan-Meier estimator for the cumulative probability of out-of-field N-site recurrences (incidence of lymph node metastases in non-irradiated LNLs, in direct analogy to the primary endpoint)
- Loco-regional control (LCR), assessed by the Kaplan-Meier estimator for the cumulative probability of recurrence of the primary tumor and/or recurrence in any neck LNL (both irradiated and non-irradiated LNLs) 2 and 3 years after finishing treatment.
- Progression-free survival (PFS), assessed by follow-up according to the study protocol. Any progression (local, regional, distant) and death of any cause will be counted as events. PFS will be reported as median (months).
- Overall survival (OS), assessed by follow-up according to the study protocol. Death of any cause will be counted as event. OS will be reported as median (months).
- Detailed patterns of failure in terms of time to event and location of the recurrence, distinguishing T-site recurrence, N-site recurrence or persistence of grossly involved lymph nodes, isolated N-site recurrences in electively treated LNLs, isolated recurrences in non-irradiated LNLs. If possible, Digital Imaging and Communications in Medicine (DICOM) data of medical imaging and radiotherapy treatment planning is collected.
- Complete blood count, including lymphocyte count. Blood examination with complete blood count, including lymphocytes taken during routine examination before first and after last radiotherapy treatment (optional). Analysis of lymphocyte count dependent on irradiated volume.
- Early (≤3 months after treatment) and late (>3 months after treatment) toxicities of treatment – overall toxicity will be recorded based on the CTCAE version 5.0 and evaluated according to the TAME methodology [28].
- Quality of life (QoL), assessed using the EORTC QLQ C30 and Head and Neck Module HN43 questionnaires at the start of treatment, as well as after completing treatment, and 6, 12 and 24 months after radiotherapy [29, 30]. QoL may be influenced by baseline characteristics, such as age and comorbidities. Therefore, participants are asked to also complete the QoL questionnaires at the start of treatment (baseline).

## Methods/Design

### Study design

The DeEscO study will be conducted as a prospective, multicenter, open label, single-arm trial at the University Hospital Zurich, the University Hospital Bern, the University Hospital Geneva, the Cantonal Hospital Aarau, the Cantonal Hospital Bellinzona, and the Réseau Hospitalier Neuchâtelois. Patients will receive treatment with definitive radiotherapy and if indicated, standard-of-care chemotherapy.

### Study population

We aim to recruit 120 oropharyngeal SCC patients referred to participating centers for definitive (chemo)radiotherapy with curative intent.

Inclusion criteria:

- Patients with a newly diagnosed (no pre-treatment) SCC of the oropharynx (i.e. tonsils, base of tongue, oropharyngeal walls, oropharyngeal surface of epiglottis; ICD-10 codes C01, C09, C10), T1-4, N0-3, irrespective of p16-status.
- Treatment with definitive (chemo)radiotherapy planned, with elective irradiation of the cervical lymph nodes.
- At the minimum 18 years of age, no upper age limit.
- ECOG performance score < 3.
- History/physical examination within 30 days prior to registration by head and neck surgeon and/or radiation oncologist and/or medical oncologist.
- FDG-PET scan prior to registration. In case of inability to perform or contra-indication, at least contrast enhanced MRI scan obligatory.
- Participants need to provide informed consent. Exclusion criteria:
- Multilevel primary tumors extending unambiguously beyond the oropharynx into the oral cavity, naso- or hypopharynx
- Distant metastases detected.
- Previous surgery, chemotherapy or radiotherapy treatment for other head and neck cancers.
- Previous surgery in head and neck region affecting the neck lymphatic system. Dissection of singular lymph nodes for diagnostic purposes before treatment start is allowed.
- Synchronous or previous malignancies. Exceptions are curatively treated basal cell carcinoma or SCC of the skin, or in situ carcinoma of the cervix uteri, low- or intermediate-risk prostate cancer or breast with a progression-free follow-up time of at least 3 years without any remaining disease burden, or other previous malignancy with a progression-free interval of at least 5 years without any remaining active/progressive disease burden regardless whether the treatment is completed or ongoing as a maintenance treatment (e.g. androgen deprivation therapy for prostate cancer).
- Pregnancy or breast feeding.
- Any severe mental or psychic disorder affecting decision making and ability to provide informed consent.

### Recruitment, screening and informed consent procedure

Recruitment of study participants, study procedures and follow-up will take place at the hospital, at one of the participating centers.

The investigators will explain to each participant the nature of the study, its purpose, the procedures involved, the expected duration, the potential risks and benefits and any discomfort it may entail. Each participant will be informed that the participation in the study is voluntary and that he or she may withdraw from the study at any time and that withdrawal of consent will not affect his or her subsequent medical assistance and treatment.

The participant will be informed that his or her medical records may be examined by authorized individuals other than their treating physician.

All participants for the study will be provided a participant information sheet and a consent form describing the study and providing sufficient information for the participant to make an informed decision about their participation in the study. Participants can decide about participation in the study until the start of treatment planning. This is usually 5-10 days after the first clinical consultation.

The formal consent of a participant, using the approved consent form, will be obtained before the participant is submitted to any study procedure.

The consent form will be signed and dated by the investigator or his designee at the same time as the participants sign. A copy of the signed informed consent will be given to the study participant. The consent form will be retained as part of the study records. The informed consent process must be documented in the patient file and any discrepancy to the process described in the protocol must be explained.

No compensation or payments will be given to the participants.

### Chemotherapy

Indications for chemotherapy are:

- Locally advanced oropharyngeal SCC (T3 or higher; or N+)
- If the patient is not eligible for treatment with Cisplatin, because of hearing loss (e.g. hearing aid), or reduced kidney function (eGFR <55 ml/min) or heart function (EF <50%), use an alternative systemic treatment as described below (options 2-4).
- In case of one or more of the following factors: age >70 years, light hearing loss, polyneuropathy, comorbidities (e.g. poorly controlled diabetes mellitus, HIV with viral load), an alternative systemic treatment as described below (options 2-4) can be considered.

Chemotherapy application (concomitant):

1. Cisplatin, 100 mg/m^2^ every three weeks

Alternatively:

2. Cisplatin, 40 mg/m^2^, weekly

If patients are not Cisplatin eligible:

1. Carboplatin AUC 2 weekly
2. Cetuximab 400 mg/m^2^ loading dose, 250 mg/m^2^ weekly

Patients receiving treatments for other unrelated illnesses have to discuss this with their treating physician, who has to record the concomitant drugs at the time of the study visit. If there is no interaction with the applied chemoradiotherapy, the concomitant drugs can be continued.

### Radiotherapy

#### Radiotherapy volumes

Radiation volumes and doses are based on Nutting et al. (Lancet Oncology, 2023) [31] and the DAHANCA 2020 guidelines [32].

The radiotherapy volumes will be defined as follows:

- GTV-T: Macroscopic gross tumor in T site, evaluated and delineated based on clinical examination and FDG-PET and/or contrast enhanced MRI.
- GTV-N: Macroscopic tumor in N site(s), evaluated and delineated based on clinical examination and FDG-PET and/or contrast enhanced MRI.
- On FDG-PET-CT, nodes with an uptake >3 SUV **<u>or</u>** size >1.5 cm **<u>or</u>** central necrosis, must be included in the GTV-N or be controlled with a negative FNA. We recommend a FNA control of all ambiguous nodes >1 cm, regardless of the SUV.
- CTV-1: Defined as a 5 mm expansion of the combined GTV (the union of GTV-T and GTV-N), corrected for anatomical barriers.
- CTV-2: Defined as a 10 mm expansion of the combined GTV, corrected for anatomical barriers.
- CTV-3: Defined as the elective target volume. LNLs are to be delineated according to Grégoire et al.[1] The LNLs to include in the CTV-3 are defined in Table 1, 2, 3, 4, and 5.
- For expansion from CTV to PTV, an expansion of 3-5 mm – to the discretion of the participating center – is used.
- All PTVs should be cropped to the skin.

**Table 1:** General Rules. If the treatment is not described in Table 2-5 or remains unclear for a patient screened for or included in this trial, the principal investigators at the University Hospital Zurich must be contacted for a treatment decision.

| General Rules |  |
| --- | --- |
| 1 | Include all involved lymph node levels (LNLs) in the CTV-3. |
| 2 | Ipsilateral level II (including IIa and IIb) is always irradiated. |
| 3 | Level Ia is never included in the CTV-3 unless clinically involved. In case of involvement of level Ia, the study PI at USZ must be contacted for a treatment decision. |
| 4 | If level IV is involved, the entire level (both IVa and IVb) is included in the CTV-3. If level IV is not involved, level IVb is never included in the CTV-3. |
| 5 | If level V is irradiated according to Table 5-7, levels Va and Vb are irradiated. Level Vc is irradiated if level Vb is involved. |
| 6 | Level VII is never included in the CTV-3 unless clinically involved. In case of level VIIa involvement, both level VIIa and VIIb are included in the CTV-3. In case of level VIIb involvement, only level VIIb (not VIIa) is included in the CTV-3. |
| 7 | If a level that is not irradiated according to Table 5-7 but partially overlaps with PTV-1, PTV-2, or PTV-3, then only the overlapping part of the level will be irradiated as part of the respective PTV. The non-overlapping part is not to be included in the CTV-3. |

If a high dose level (CTV-1/CTV-2) extends into a LNL that is not included in the elective nodal volume (CTV-3) defined according to Table 1-5, only this extension part will be included in the radiation volume and not the complete LNL.

Nodal levels must be delineated according to Gregoire et al., Radiother Oncol, 2014 [1], and organs at risk according to Brouwer et al., Radiother Oncol, 2015 [33]. The pharyngeal constrictor muscle is contoured as three structures: superior, middle and inferior; for the constraints the superior and middle pharyngeal constrictor muscles are considered a single structure.

#### Radiotherapy doses

The following radiotherapy doses will be applied to the target volumes:

- CTV-1/PTV-1: 65-70 Gy*
- CTV-2/PTV-2: 60-65 Gy*
- CTV-3/PTV-3: 50-54 Gy*

*Radiotherapy will be given as simultaneous integrated boost (SIB) in 30-35 fractions of 1.8-2.2 Gy (in CTV-1 and CTV-2) and 1.47-1.8 Gy (in CTV-3) over 6-7 weeks – to discretion of the treating participating center.

PTV-1 and PTV-2 may receive the same radiotherapy dose (as described e.g. in Nutting et al. – in this case there would be no “intermediate dose”/ “intermediate risk volume”).

Patients not receiving systemic treatment for any reason are allowed to be treated with accelerated radiotherapy, 6x per week.

#### Personalized elective nodal CTV de-escalation

Regarding the volume de-escalation, Table 2, 3, 4, and 5 describe the elective treatment volume (CTV-3), dependent on T-stage, midline extension, and clinically involved levels.

**Table 2.** Treatment recommendations for central tumors or tumors without clear lateralization. *Only irradiate level IB electively if level IIA on the same side is involved.

|  |  |  | To be included in CTV-N | To be included in CTV-N |
| --- | --- | --- | --- | --- |
| T-stage | Nodal involvement side one | Nodal involvement side two | Side one neck | Side two neck |
| T1-2 | - | - | II | II |
| T1-2 | II | - | II, III | II, III |
| T1-2 | III | - | II, III, IVa | II, III |
| T1-2 | II, III | - | II, III, IVa | II, III |
| T1-2 | II | II | (Ib)*, II, III | (Ib)*, II, III |
| T1-2 | II | III | (Ib)*, II, III | II, III, IVa |
| T1-2 | III | III | II, III, IVa | II, III, IVa |
| T1-2 | II, III | II | (Ib)*, II, III, IVa | II, III |
| T1-2 | II, III | III | (Ib)*, II, III, IVa | II, III, IVa |
| T1-2 | II, III | II, III | (Ib)*, II, III, IVa | (Ib)*, II, III, IVa |
| T3-4 | - | - | II, III | II, III |
| T3-4 | II | - | II, III | II, III |
| T3-4 | III | - | II, III, IVa | II, III |
| T3-4 | II | II | (Ib)*, II, III | (Ib)*, II, III |
| T3-4 | II | III | (Ib)*, II, III | II, III, IVa |
| T3-4 | III | III | II, III, IVa | II, III, IVa |
| T3-4 | II, III | - | (Ib)*, II, III, IVa | II, III |
| T3-4 | II, III | II | (Ib)*, II, III, IVa | (Ib)*, II, III |
| T3-4 | II, III | III | (Ib)*, II, III, IVa | II, III, IVa |
| T3-4 | II, III | II, III | (Ib)*, II, III, IVa | (Ib)*, II, III, IVa |

**Table 3.** Treatment recommendations for lateralized tumors, with midline extension and early T-stage (T1-2). * Only irradiate level IB electively if level IIA on the same side is involved. ** Level Va and Vb are irradiated.

|  |  | To be included in CTV-N | To be included in CTV-N |
| --- | --- | --- | --- |
| Nodal involvement ipsilateral | Nodal involvement contralateral | Ipsilateral Neck | Contralateral Neck |
| - | - | II | II |
| II | - | II, III | II |
| III | - | II, III, IVa | II |
| VII | - | II, III, VII | II |
| II, III | - | II, III, IVa | II |
| II, VII | - | II, III, VII | II |
| III, VII | - | II, III, IVa, VII | II |
| II, III, IV | - | II, III, IV, V** | II |
| II | II | II, III | II, III |
| II, III | II | II, III, IVa | II, III |
| II, III, IV | II | II, III, IV, V** | II, III |
| II, III | II, III | (Ib)*, II, III, IVa | II, III, IVa |

**Table 4.** Treatment recommendations for lateralized tumors, with midline extension and advanced T-stage (T3-4). * Only irradiate level IB electively if level IIA on the same side is involved. ** Level Va and Vb are irradiated. Level Vc is irradiated if level Vb is involved.

|  |  | To be included in CTV-N | To be included in CTV-N |
| --- | --- | --- | --- |
| Nodal involvement ipsilateral | Nodal involvement contralateral | Ipsilateral Neck | Contralateral Neck |
| - | - | II, III | II |
| II | - | II, III | II |
| III | - | II, III, IVa | II |
| II, III | - | II, III, IVa | II |
| II, VII | - | II, III, VII | II |
| Ib, II, III | - | Ib, II, III, IVa | II |
| II, III, IV | - | II, III, IV, V** | II |
| II, III, IV, V | - | (Ib)*, II, III, IV, V** | II |
| II | II | (Ib)*, II, III | II, III |
| II, III | II | (Ib)*, II, III, IVa | II, III |
| II, VII | II | II, III, VII | II, III |
| Ib, II, III | II | Ib, II, III, IVa | II, III |
| II, III, IV | II | II, III, IV, V** | II, III |
| II, III, VII | II | II, III, IVa, VII | II, III |
| II | II, III | (Ib)*, II, III | II, III, IVa |
| II, III, IV | II, III | II, III, IVa, V** | II, III, IVa |
| II, III | II, III | (Ib)*, II, III, IVa | II, III, IVa |
| II, III | II, III, IV | (Ib)*, II, III, IVa | II, III, IV, V** |

**Table 5.** Treatment recommendations for lateralized tumors, without midline extension and early T-stage (T1-2). * Only irradiate level IB electively if level IIA on the same side is involved. ** Level Va and Vb are irradiated. Level Vc is irradiated if level Vb is involved.

|  |  | To be included in CTV-N | To be included in CTV-N |
| --- | --- | --- | --- |
| Nodal involvement ipsilateral | Nodal involvement contralateral | Ipsilateral Neck | Contralateral Neck |
| - | - | II | - |
| II | - | II, III | - |
| III | - | II, III, IVa | - |
| IV | - | II, III, IV, V** | - |
| V | - | II, III, IVa, V** | - |
| Ib, II | - | Ib, II, III | - |
| II, III | - | II, III, IVa | - |
| II, V | - | II, III, IVa, V** | - |
| II, VII | - | II, III, VII | - |
| Ib, II, III | - | Ib, II, III, IVa | - |
| II, III, IV | - | II, III, IV, V** | - |
| II, III, V | - | II, III, IVa, V** | - |
| II, III, VII | - | II, III, IVa, VII | - |
| II, III, IV, V | - | II, III, IV, V** | - |
| II | II | II, III | II, III |
| II, III | II | (Ib)*, II, III, IVa | II, III |
| II, III, IV | II | II, III, IV, V** | II, III |

For the stratification of patients into Table 2-5, the following definitions apply:

- “With midline extension” is defined as a patient in whom any part of the primary tumor GTV (GTV-T) extends beyond the midsagittal plane.
- “Central tumors” are those with similar tumor extension on both sides of the midsagittal plane by the judgement of the treating physician. Tumors extending on both sides but with the main tumor mass located on one side are instead classified as “lateralized tumors with midline extension”.
- “Early T-stage” is defined as T-stages 1-2.
- “Advanced T-stage” is defined as T-stages 3-4.

For the definition of clinical lymph node involvement per level, the following rules apply:

- On FDG-PET-CT, nodes with an uptake >3 SUV or size >1.5 cm or central necrosis are to be considered clinically involved.
- In case of ambiguity after radiological work-up regarding lymph node involvement, FNA is recommended.
- In case of a lymph node conglomerate or large nodes extending over 2 levels, both levels should be considered as involved.

Figure 2 A-F provide schematic examples of the personalized elective nodal CTV de-escalation.

**Figure 1:**
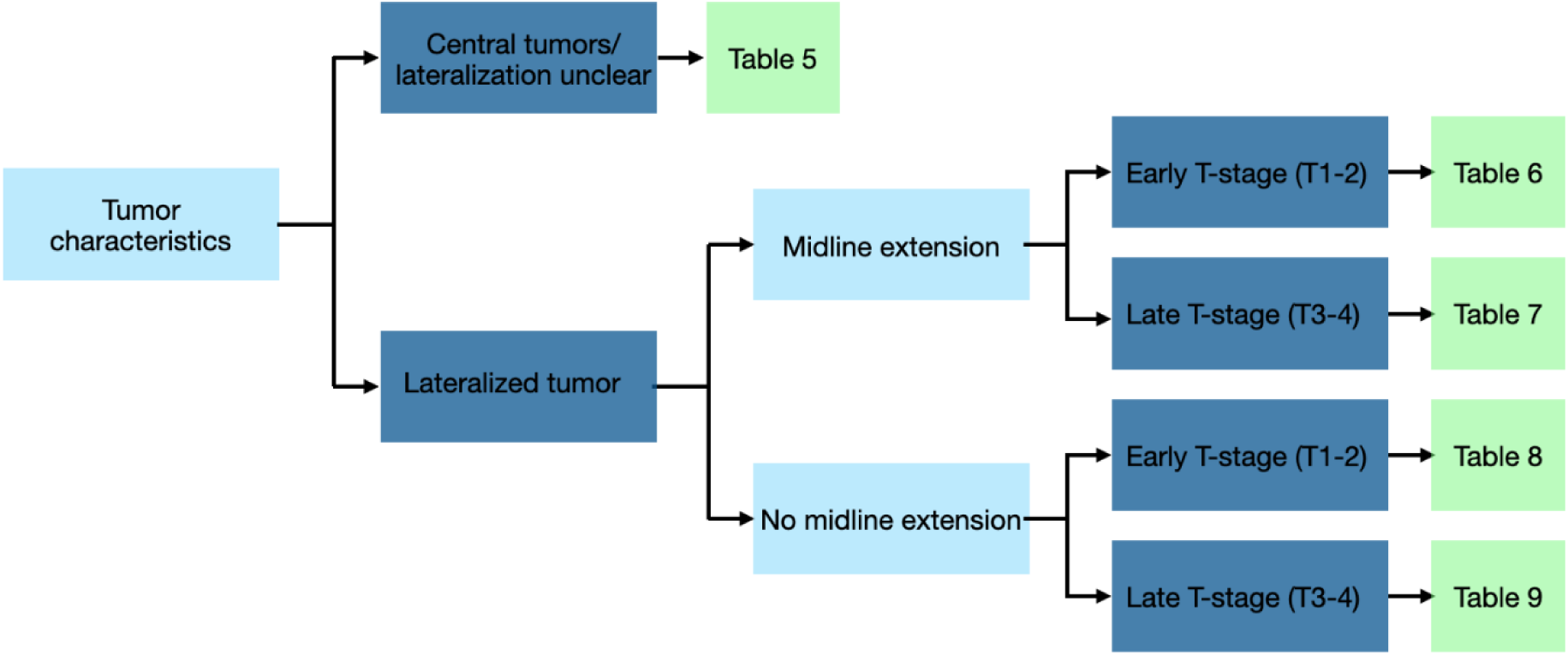
*Decision tree to choose the table for looking up the personalized treatment.* Figure 1 provides a decision tree on which table to use to determine the personalized treatment based on individual tumor characteristics. The tables cover the most commonly observed cases in our database (85% of the cases in our database) and a select number of plausible cases. The investigators at University Hospital Zurich have a table with all (theoretically) possible cases and their treatment recommendations available. The general rules in Table 1 are applied.

**Figures 2.**
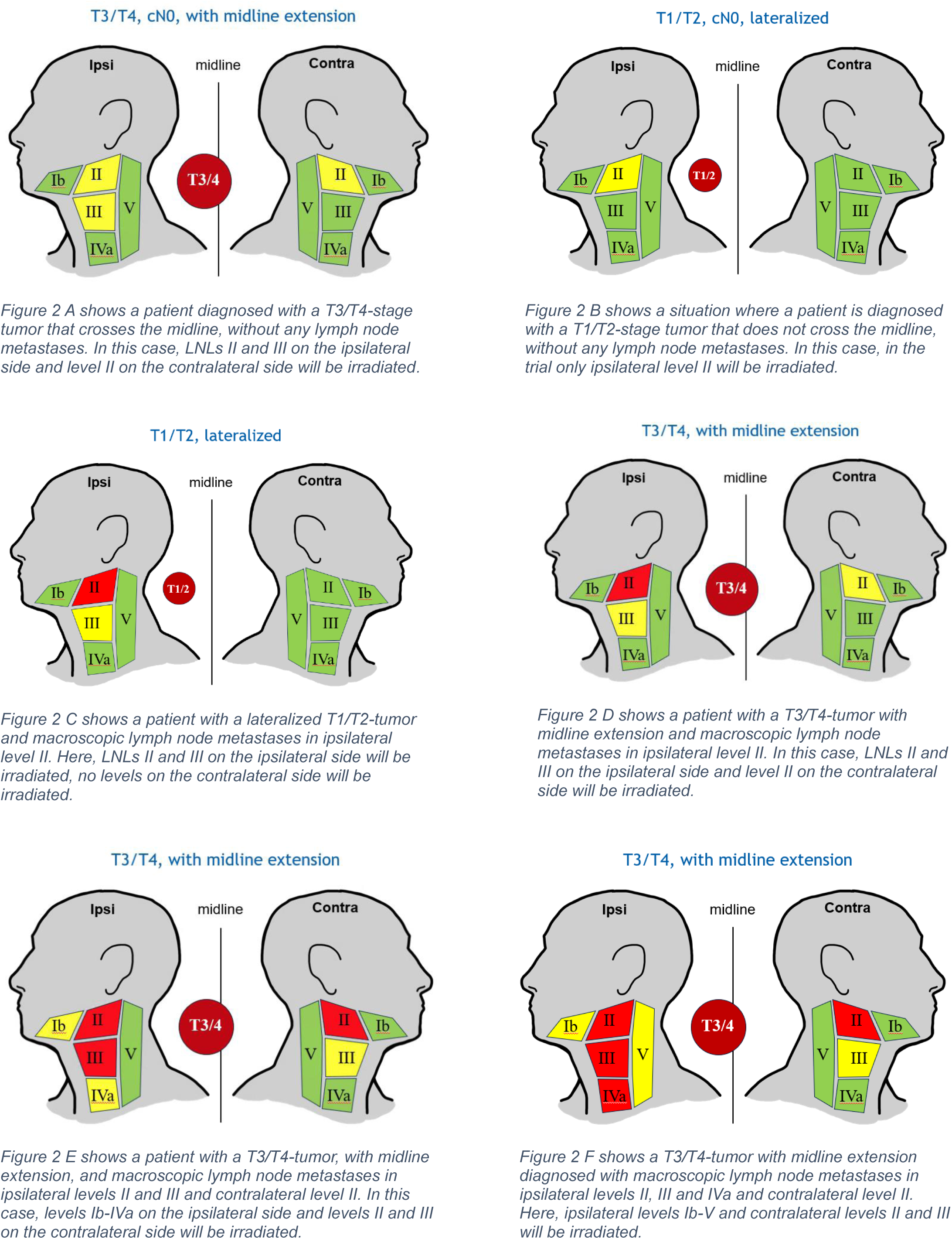
A-F. Schematic examples of the personalized elective nodal CTV de-escalation.

### Follow-up

Table 6 and 7 show a schematic overview of the schedule of assessment during treatment and during the follow-up. During the treatment, there will be weekly clinical visits to evaluate toxicities.

**Table 6.** Schedule of Assessments – During Treatment. * Dependent on treatment schedule, the (chemo)radiotherapy treatment is applied in 6 or 7 weeks. Participants are requested to complete the QoL questionnaires after finishing the (chemo)radiotherapy treatment. ** During routine clinical visit. Additional procedure and optional in case patients receive no chemotherapy (total blood count for secondary endpoint)

| Time (week) (+/- 5 days) | Before first radiotherapy treatment (-2 weeks) | +1 | +2 | +3 | +4 | +5 | +6 | (+7)* | After last radiotherapy treatment (+1 week) |
| --- | --- | --- | --- | --- | --- | --- | --- | --- | --- |
| Visit |  | 2 | 3 | 4 | 5 | 6 | 7 | (8) |  |
| Physical examination |  | ✓ | ✓ | ✓ | ✓ | ✓ | ✓ | ✓ |  |
| Laboratory (optional)** | + / ✓ |  |  |  |  |  |  |  | + / ✓ |
| (Chemo)radiotherapy treatment |  | ✓ | ✓ | ✓ | ✓ | ✓ | ✓ | ✓ |  |
| Questionnaire (QLQ-C30 and QLQ-H&N43) | + |  |  |  |  |  |  |  | + |
| Toxicity evaluation (CTCAE v5.0) |  | ✓ | ✓ | ✓ | ✓ | ✓ | ✓ | ✓ |  |

**Table 7.** Schedule of Assessments – Follow-up. *PET-CT is recommended in case of no contra-indications.

| Time (week/month) | +6-8 weeks (+/- 10 days) | +3-4 months (+/- 2 weeks) | +6 months (+/- 1 month) | +9 months (+/- 1 month) | +12 months (+/- 1 month) |
| --- | --- | --- | --- | --- | --- |
| Visit | 9 | 10 | 11 | 12 | 13 |
| Physical examination | ✓ | ✓ | ✓ | ✓ | ✓ |
| Ultrasound | ✓ |  | ✓<br>(if no other imaging performed) |  | ✓<br>(if no other imaging performed) |
| Follow-up with imaging |  | ✓<br>(PET-CT or enhanced contrast MRI)* | If indicated | ✓<br>(PET-CT or enhanced contrast MRI)* | If indicated |
| Questionnaire (QLQ-C30 and QLQ-H&N43) |  |  | + |  | + |
| Toxicity evaluation (CTCAE v5.0) | ✓ | ✓ | ✓ | ✓ | ✓ |
| Time (month) | +15 months (+/- 1 month) | +18 months (+/- 1 month) | +21 months (+/- 1 month) | +24 months (+/- 1 month) | +30 months (+/- 1 month) |
| Visit | 14 | 15 | 16 | 17 | 18 |
| Physical examination | ✓ | ✓ | ✓ | ✓ | ✓ |
| Ultrasound | ✓<br>(if no other imaging performed) | ✓<br>(if no other imaging performed) | ✓<br>(if no other imaging performed) |  | ✓<br>(if no other imaging performed) |
| Follow-up with imaging | If indicated | If indicated | If indicated | ✓<br>(PET-CT or enhanced contrast MRI)* | If indicated |
| Questionnaire (QLQ-C30 and QLQ-H&N43) |  |  |  | + |  |
| Toxicity evaluation (CTCAE v5.0) | ✓ | ✓ | ✓ | ✓ | ✓ |
| Time (month) | +36 months (+/- 1 month) |  |  |  |  |
| Visit | 19 |  |  |  |  |
| Physical examination | ✓ |  |  |  |  |
| Ultrasound | ✓<br>(if no other imaging performed) |  |  |  |  |
| Follow-up with imaging | If indicated |  |  |  |  |
| Questionnaire (QLQ-C30 and QLQ-H&N43) |  |  |  |  |  |
| Toxicity evaluation (CTCAE v5.0) | ✓ |  |  |  |  |

After finishing treatment:

- The first clinical assessment will take place after 6-8 weeks. Early progression should be ruled out and toxicity should be recorded. An ultrasound examination of the neck LNLs is mandatory. Only nodes >1 cm not present at baseline should be cytologically controlled with fine-needle aspiration (FNA).
- The first (mandatory) follow-up PET-CT scan will take place 3-4 months after finishing treatment. If there are contra-indications for a PET-CT scan, follow-up with at least a contrast enhanced MRI must take place.
- Afterwards, follow-up visits will take place every 3 months during the first 2 years of follow-up, including assessment of toxicities and imaging if needed. If no imaging is performed, an ultrasound examination is mandatory to rule out lymph node recurrences/progression.
- Further (mandatory) standard follow-up PET-CT scans will take place 9-10 months and 2 years after finishing treatment.
- During the third year of follow-up, follow-up visits will take place every 6 months. If no imaging is performed, an ultrasound examination is mandatory to rule out lymph node recurrences/progression.

On PET-CT scan newly occurred suspicious nodes with an uptake >3 SUV or size >1.5 cm or central necrosis should always be controlled with FNA or treated with a selective salvage procedure.

Suspicious lymph nodes already present at baseline and treated as GTV, that are still present in the follow-up PET-CT scan with an uptake of >3 SUV, can be monitored with subsequent imaging every 3 months as long as the lymph nodes are decreasing in size or uptake, or alternatively the suspicious nodes can be treated with a selective salvage neck dissection. Unambiguous positive nodes, with e.g. positive FNA results or progression signs in subsequent imaging should be always treated with salvage elective neck dissection if possible.

Every newly diagnosed nodal metastasis (i.e. not present at primary diagnosis) should be recorded, including level(s) of occurrence and FNA applied. Only metastases not included in the irradiated volume (CTV) will be counted as events towards the primary endpoint. Marginal miss, i.e. new occurring lymph node outside of CTV-3, including ambiguous cases, but inside the 50% prescription isodose of PTV-3, is also counted as event towards the primary endpoint.

### Serious adverse events

All serious adverse events (SAEs) are documented. Only related SAEs are documented and reported immediately (within 24 hours) to the Sponsor-Investigator of the study. If it cannot be excluded that the SAE occurring is attributable to the intervention under investigation, the Sponsor-Investigator reports it to the Ethics Committee via BASEC within 15 days. A planned hospitalization for pre-existing condition, or a procedure required by the protocol, without a serious deterioration in health is not considered to be a SAE.

### Statistical methods

The primary goal of this study is to determine the rate of out-of-field N-site recurrences in nonirradiated lymph node levels after two years. The intervention is designed such that we expect a rate of out-of-field N-site recurrences in non-irradiated lymph node levels of <10% (resulting in a value of 0.9 of the survival function at two years). The goal is to measure the cumulative probability of out-of-field recurrence with a precision resulting in a width of 10% of the 90% confidence interval of the Kaplan-Meier estimator at 2 years).

#### Sample size calculation

120 patients will be included in this study. Under the assumption of no censoring or drop-outs, the intended width of 10% for the 90% confidence interval (from 0.84 to 0.94) for the survival probability of 0.9 can be obtained with 104 patients. By including 120 patients, we account for an amount of censoring or dropouts of 13%. The confidence interval was calculated for the log-minus-log transformation of the survival function using the asymptotic variance approximation as described in Nagashima et al. [34] and then back transformed. In order to achieve adequate participant enrolment, six centers in Switzerland participate in the trial.

#### Statistical analyses

The primary endpoint will be evaluated according to the per-protocol treatment, although all primary analyses will follow both the per-protocol and intention-to-treat principles.

For the per-protocol analysis, we will only include patients who completed the treatment as described in the study procedures. For the intention-to-treat analysis, all participants included in the study who provided informed consent, will be analyzed for the primary endpoint (out-of-field lymph node recurrence) – regardless of any treatment changes afterwards.

The primary endpoint “Out-of-field lymph node recurrence” will be reported as the Kaplan-Meier estimator for the time-to-event at 2 years. The calculation of the Kaplan-Meier estimator at 2 years will include the events detected at the 24-months follow-up visit (which may occur at 2 years +/- 1 month). Patients lost to follow-up and patients who die due to any cause are right-censored.

In addition to the overall rate of Out-of-field lymph node recurrences, we will analyze whether 1) Out-of-field lymph node recurrence was the initial cause for loco-regional failure, or whether 2) recurrence/persistence of the primary tumor or grossly involved lymph nodes present at baseline was observed simultaneously or prior to the Out-of-field lymph node recurrence. In case 1, irradiating the respective LNL electively may have prevented loco-regional failure. In case 2, loco-regional failure may have occurred irrespective of volume-de-escalation, and the Out-of-field lymph node recurrence may be due to lymphatic spread of the recurrent/persistent tumor. To determine the rate of "Out-of-field lymph node recurrences being the initial cause for loco-regional failure", we will perform Kaplan-Meier analysis where patients with recurrence/persistence of the primary tumor or grossly involved lymph nodes present at baseline are right-censored.

Dropout will be dealt with as independent right censoring. Participants who provided informed consent will be included in the ITT-analysis. Only patients who discontinued the study due to consent withdrawal will be replaced with new patients and should get only standard oncological follow-up and no further study-specific follow up. All other patients who discontinue the study due to intervention associated reasons or (chemo)radiotherapy/disease-associated reasons will be included in the ITT-analysis and receive study-specific follow up.

### Safety considerations and rules for early termination

SAEs – both related and unrelated to the (chemo)radiotherapy treatment – will be monitored and recorded. In addition, we will perform a safety interim analysis when 60 patients have been enrolled. The following procedure will be followed.

1. The patients for whom the 6-month follow-up visit has been completed and reported are identified. Let N be the number of patients with at least 6 months follow-up.
2. If N is smaller than 20, recruitment of further patients will be paused until the 6-month follow-up visit is reported for at least 20 patients.
3. For the N patients with at least 6 months follow-up, the number of events towards the primary endpoint will be counted, denoted by k.
4. The number of events will be compared to a binomial distribution with parameters N and p=0.1. If the probability to observe k or more events among N patients assuming an event probability of 10% is less than 10%, recruitment of further patients will be paused for 6 months. Otherwise, the study will be continued.
5. If the study is paused, the analysis will be repeated after 6 months, i.e. the patients who completed the 6-month follow-up visit are considered (N) and the number of events towards the primary endpoint is determined (k). If the probability to observe k or more events is less than 5%, assuming a binomial distribution with parameters N and p=0.1, the study will be terminated (i.e. no further patients will be enrolled). Otherwise, the study will be continued.

As an additional termination criterion, the total number of events towards the primary endpoint (out-of-field N-site recurrences) will be monitored. If at any point in time during recruitment, the total number of events reported to the sponsor exceeds 20, no additional patients will be enrolled.

The stop for futility/safety does not increase the type I error and thus no adjustments of the confidence interval are necessary.

### Insurance

Insurance for all participating centers is covered by “Versicherung für klinische Versuche und nichtklinische Versuche“ by Zürich Versicherungs-Gesellschaft AG (Policy no.: 14.970.888). The insurance (guarantee) must cover damage occurring up to 20 years after the end of the clinical trial.

Any damage developed in relation to study participation is covered by this insurance. So as not to forfeit their insurance cover, the participants themselves must strictly follow the instructions of the study personnel. Participants must not be involved in any other medical treatment without the permission of the principal investigator (emergency excluded). Medical emergency treatment must be reported immediately to the investigator. The investigator must also be informed instantly, in the event of health problems or other damages during or after the course of study treatment.

The investigator will allow delegates of the insurance company to have access to the source data/documents as necessary to clarify a case of damage related to study participation. All parties involved will keep the patient data strictly confidential.

A copy of the insurance certificate will be placed in the Investigator’s Site File.

## Data collection and management

### Data collection and storage

The study will strictly follow the protocol. If any changes become necessary, they must be laid down in an amendment to the protocol. All amendments of the protocol must be signed by the Sponsor-Investigator and local Principal Investigators and if essential submitted to CEC.

All study data will be entered into REDCap; an electronic case report form database. In addition, imaging data (DICOM) including the radiotherapy plan with structure and dose information and the PET-scans up to the end of the follow-up period will be collected, pseudonymized in all centers and shared with the sponsor via USZ-data-transfer. Pseudonymized study data will be securely stored in REDCap, pseudonymized medical imaging and treatment plans will be stored on a secured server in the University Hospital of Zurich, Switzerland.

Study participants can choose to fill out the QoL questionnaires online or on paper. In case participants want to complete the questionnaires online, we will store their e-mail address in our secured database. The e-mail address is marked as identifier in the database and can neither be accessed nor exported by the Sponsor. It will therefore not appear in the study data. Only specific roles (i.e. data manager, project manager, monitor) can view all data in the database for the performance of their tasks, but they underly strict confidentiality. Participants’ e-mail addresses will only be used for the purpose of automatically sending the QoL questionnaires in electronic form. The patients are informed about this procedure and agree to or decline in the ICF the use of their e-mail address.

### Confidentiality and safe handling of data

Data generation, transmission, storage and analysis of personal data within this project strictly follow Swiss legal requirements for data protection.

### Case Report Forms

The eCRF will capture all planned visits and examinations, as well as all parameters and data described in the study protocol. No detail described/not described in the protocol will be omitted/added in the eCRF. Wording and expressions used in the protocol (e.g.: inclusion and exclusion criteria) will be used identically in the eCRF.

The investigators will use electronic case report forms (eCRF), one for each enrolled study participant, to be filled in with all relevant data pertaining to the participant during the study. All participants who either entered the study or were considered not eligible or were eligible but not enrolled into the study additionally have to be documented on a screening log. The investigator will document the participation of each study participant on the Enrolment Log.

For data and query management, monitoring, reporting and coding, the internet-based secure database REDCap® developed in agreement to the Good Clinical Practice (GCP) guidelines provided by the Clinical Trials Centre (CTC) Zurich will be used for this study. It is the responsibility of the investigator to assure that all data in the course of the study will be entered completely and correctly in the respective database. Corrections in the eCRF may only be done by the investigator or by other authorized persons. In case of corrections the original data entries will be archived in the system and can be made visible. For all data entries and corrections date, time of day and person who is performing the entries will be generated automatically.

eCRFs will be kept current to reflect participant status at each phase during the course of study. Participants must not to be identified in the eCRF by name. Appropriate coded identification (e.g. Participant Number) will be used.

Any authorized person, who may perform data entries and changes in the eCRF, will be identifiable and clearly member of the study team according to the delegation log. A list with signatures and initials of all authorized persons will be filed in the study site file and the trial master file, respectively.

Documented medical histories and narrative statements relative to the participants’ progress during the study will be maintained. These records will also include the following: originals or copies of laboratory and other medical test results (e.g. ECGs, etc.) which must be kept on file with the individual participant’s eCRF.

Patient medical history, concomitant medication, results of physical examination, pathology and laboratory results, (chemo)radiotherapy prescription, and correspondence with other treating physicians will be maintained for each participant as in normal clinical routine in the clinic electronic patient file.

The investigators assure to perform a complete and accurate documentation of the participant data in the eCRF. All data entered into the eCRF will also be available in the individual participant file either as printouts or as notes taken by either the investigator or another responsible person assigned by the investigator.

### Confidentiality and coding

Direct access to source documents will be permitted for the purposes of monitoring, audits and inspections.

Trial and participant data will be handled with the utmost discretion and is only accessible to authorized personnel who require the data to fulfil their duties within the scope of the study. On the CRFs and other study specific documents (including medical imaging, DICOM), participants are only identified by a unique participant number and data will be pseudonymized. Only a few people directly involved in the trial will have access to the pseudonymization-key and only in order to perform assessments and analyses, which stand in direct relation to the present study.

The participant identification list will be stored by the person responsible at the participating centers. A safety back-up in REDCap will be stored, to protect the data from accidental alteration and deletion. A quality assurance audit/inspection may be conducted. Only non-genetic data are used.

### Retention and destruction of study data and imaging

No biological material will be acquired exclusively for study purposes.

All patient files and source data will be archived for the longest possible period according to the feasibility of the investigational site, e.g. hospital, institution or private practice.

All study data will be archived in the study office of the clinic for Radiation Oncology of the Zurich University Hospital for a minimum of 20 years after study termination or premature termination of the clinical trial (KlinV Art. 45). The investigators have to archive major administrative documents (correspondence with ethical committee, authorities, sponsor, etc.), the patient identification log, the signed informed consent forms, and the main study documents (protocol, amendments) for the same period. The original patient records have to be archived according to the standard procedures of the respective institution.

The data may be used in the future for research that is not yet defined (“further use”). The time for data storage for potential further use projects is not defined.

### Audits and Inspections

A quality assurance audit/inspection of this study may be conducted by the CEC. The quality assurance auditor/inspector will have access to all medical records, the investigator’s study related files and correspondence, and the informed consent documentation that is relevant to this clinical study.

The investigator will allow the people responsible for the audit or the inspection to have access to the source data/documents and to answer any questions arising. All parties involved will keep the patient data strictly confidential.

### Monitoring

External, independent monitoring will be performed by University Hospital Zurich, Clinical Trials Center. The monitoring strategy will be a low-risk approach.

A quality assurance audit/inspection of this study may be conducted by the CEC. The quality assurance auditor/inspector will have access to all medical records, the investigator’s study related files and correspondence, and the informed consent documentation that is relevant to this clinical study.

The investigator will allow the people responsible for the audit or the inspection to have access to the source data/documents and to answer any questions arising. All parties involved will keep the patient data strictly confidential.

## Discussion

Elective irradiation of the neck lymph node drainage in patients with oropharyngeal cancer is associated with multiple short- and long-term side effects such as pain, dysphagia, and lymphedema, negatively affecting QoL. Yet, de-escalation of treatment has not been proven safe enough to implement in clinical practice thus far. To our knowledge, the DeEscO clinical trial is the first trial where the de-escalation strategy is personalized based on clinical factors and calculations of occult metastases risks using a statistical model.

The elective irradiation of the neck LNLs is associated with early and late toxicity. Toxicities such as pain, dermatitis, mucositis, but also long-term sequela like swallowing dysfunction, lymphedema and dysgeusia are commonly described, which potentially lead to hospitalization or long-term symptoms with subsequent life-quality impairment [6, 7]. In addition, patients receiving combined treatment with chemoradiotherapy reported high levels of depressive symptomatology [35]. In the DeEscO trial, we include toxicities graded according to CTCAE version 5 and patient-reported outcome measures (PROMs) using quality of life questionnaires as secondary endpoints. Additionally, as a secondary outcome measure, we will analyze lymphocyte counts in patients treated with different areas of elective CTV to evaluate possible immune-sparing effects of radiotherapy treatment de-escalation.

Multiple studies regarding primary and adjuvant treatment of oropharyngeal SCC have evaluated potential ways to de-escalate treatment and reduce toxicity, such as dose reduction or change of chemotherapeutic agent. Several studies showed favorable oncological and toxicity outcomes [10, 36–43]. Omission of the contralateral LNLs in carefully selected patients with HPV-positive oropharyngeal SCC has been shown to be safe and effective in retrospective studies [44, 45]. However, other studies, including a meta-analysis by Petrelli et al. for HPV+ oropharyngeal SCC including studies with various de-escalation strategies (surgery vs surgery plus adjuvant radiotherapy, radiotherapy vs chemoradiotherapy, cetuximab vs cisplatin, hypofractionated radiotherapy, normal fractionation vs acceleration, dose reduction), showed a reduction in overall survival (OS), progression-free survival (PFS), loco-regional and distant disease control for de-escalated treatments – concluding that de-escalation cannot (yet) be safely adopted in clinical practice [46, 47]. Another possible de-escalation strategy, which is pursued here, is to reduce the elective clinical target volume. Few studies have been published on volume de-escalation, and results regarding loco-regional control and toxicity reduction seem promising [48]. The study of Martínez Carrillo included postoperative patients with oral cavity or oropharyngeal SCC. The elective CTV was selected based on clinicopathological factors, such as midline extension, the number of nodes involved and the dissected levels. Apart from the dissected levels, these factors are the same as in the DeEscO trial. The irradiated volume as part of the elective CTV appears to be similar to the volumes defined in DeEscO, e.g. by including the LNLs adjacent to an involved LNL. Nodal recurrence was most frequently detected in high-risk nodal levels receiving high irradiation doses [48]. In addition, currently running trials such as the EVADER Trial by the Canadian Cancer Trials Group (CCTG) (clinicaltrials.gov NCT03822897) apply similar volume de-escalation strategies. All patients are treated with a volume-reduced elective CTV, with the extent of elective CTV determined by the subsite of the primary tumor within the oropharynx and by the number and distribution of involved neck lymph nodes. Only HPV positive patients with oropharyngeal SCC, T1-3, N0-1 are included in the EVADER trial. In the DeEscO trial we use similar factors to determine the elective CTV, such as T-stage, midline extension, and the involvement of LNLs. Our calculations do however not include the subsite of the primary tumor and the DeEscO trial is not restricted to HPV positive patients. The DeEscO trial provides a more personalized treatment approach than the EVADER trial, as the elective CTV is based on the specific macroscopic involved LNLs – which levels are involved and how many levels are involved - compared to N0 or N1 categories used in the EVADER trial. Another similar currently running trial aiming to achieve lymphocyte sparing is the Lymphocyte-Sparing Artificial Intelligence-guided Radio-Immunotherapy (LySAIRI) Trial. Here, a reduction in the elective nodal CTV irradiation volume is combined with trans retinoic acid for immunomodulation [49]. The SELECT-trial makes use of lymphatic mapping using SPECT-CT to guide elective contralateral neck treatment. In this trial, de-escalated treatment of the contralateral side using lymphatic mapping is compared to bilateral neck irradiation [50].

Recently, other efforts regarding personalized de-escalation, for example based on radiologic features such as treatment hypoxia, showed promising results regarding personalizing treatment dose and herewith reducing treatment-associated toxicity [51]. In the REWRITE Trial, radiotherapy was only delivered to the primary tumor, involved lymph nodes, and the nodal level immediately adjacent to the primary tumor and/or involved lymph nodes in patients with SCC head and neck cancers. The trial showed that in frail patients very limited N0 neck irradiation was associated with a very low probability of nodal relapse in the non-irradiated neck [52]. In the DeEscO clinical trial, the de-escalation is based on a statistical model developed from an international multicentric database with 598 oropharyngeal SCC patients. Thus, the individualized risk for lymph node metastases can be determined separately for each patient, resulting in a personalized treatment based on a patient’s individual characteristics and clinical nodal involvement. To our knowledge, there have been no previous attempts for such a tailored treatment de-escalation in head and neck cancers.

The DeEscO trial will provide information on the efficacy and safety of personalized reduction of the elective nodal CTV in oropharyngeal SCC patients, based on the individual patient’s state of disease progression and risk factors. We are convinced that our approach could help improve the current standard of care for patients diagnosed with oropharyngeal SCC.

## Study status

The first center opened in January 2025. Three other centers opened in February 2025. The other 2 centers are planned to follow shortly afterwards. Recruitment is estimated to be completed by January 2027.

## Data Availability

All data leading to this trial data has been made publicly available.
Anonymized data produced by the presented ongoing clinical trial will be made available after the study has concluded.

## List of abbreviations

BASEC: Business Administration System for Ethical Committees
CRF: Case Report Form
CT: Computed Tomography
CTCAE: Common Terminology Criteria for Adverse Events
CTV: Clinical Target Volume
DICOM: Digital Imaging and Communications in Medicine
ECOG: Eastern Cooperative Oncology Group
eCRF: electronic Case Report Form
FDG: Fluorodeoxyglucose
FNA: Fine Needle Aspiration
GCP: Good Clinical Practice
GTV: Gross Tumor Volume
GTV-T: Macroscopic Gross Tumor Volume in T site
GTV-N: Macroscopic Gross Tumor Volume in N site(s)
Gy: Gray
HMM: Hidden Markov Models
HRA: Human Research Act (in German: HFG, in French: LRH, in Italian: LRUm)
ITT: Intention-To-Treat analysis
LCR: Locoregional Control Rate
LNL: Lymph Node Level
MRI: Magnetic Resonance Imaging
OAR: Organs At Risk
OS: Overall Survival
PET-CT: Positron Emission Tomography and Computed Tomography
PFS: Progression-Free Survival
PROM: Patient-reported outcome measure
PTV: Planning Target Volume
QoL: Quality of Life
SAE: Serious Adverse Event
SCC: Squamous Cell Carcinoma
SUV: Standardized Uptake Value

## Declarations

### Ethics approval and consent to participate

Ethical approval was obtained on 21/01/2025, BASEC-Nr 2024-01936. The study will be performed in accordance with the Declaration of Helsinki, the principles of Good Clinical Practice, the Human Research Act (HRA) and the Human Research Ordinance (HRO), as well as other relevant local regulations. Written informed consent is obtained from each participant prior to their inclusion in the study.

Protocol amendments will be communicated in written form to the participating centers.

### Consent for publication

Written informed consent is obtained from each participant prior to their inclusion in the study.

### Availability of data and materials

The previously collected multi-institutional dataset of 598 oropharyngeal SCC patients in whom the detailed patterns of lymph node involvement are reported is publicly available on www.LyProX.org. The data collected during the clinical trial will be published and be publicly available, after evaluation of each endpoint and concurrently with respective publications.

### Competing interests

The authors declare no competing interests.

### Funding

The clinical trial will be funded internally. Preparation of the protocol and research leading to the design of the clinical trial was supported by the Clinical research priority program Artificial intelligence in oncological imaging of the University Zürich, and by the Swiss cancer research foundation under grant KFS-5645-08-2022.

### Trial Sponsor

Prof. Dr. med. Panagiotis Balermpas

Department of Radiation Oncology University Hospital Zurich Raemistrasse 100 CH 8091 ZURICH Switzerland E-Mail

### Authors’ contributions

P.B. is the principal investigator and sponsor. E.L., Y.P.H., R.L., J.U. and P.B. contributed to the conception and design of the study. E.L. drafted the initial manuscript. Y.P.H., R.L., G.M., M.G., D.F., S.B., O.E., R.G., S.T., F.M., S.M., O.R., J.U. and P.B. reviewed and contributed to the manuscript. All authors read and approved the final manuscript.

## Acknowledgements

We would like to thank the study coordination offices and their staff at all participating centers for their support with this project.

## Notes

### Competing Interest Statement

The authors have declared no competing interest.

### Clinical Trial

NCT03822897

### Funding Statement

This clinical trial will be funded internally. Preparation of the protocol and research leading to the design of the clinical
trial was supported by the Clinical research priority program Artificial intelligence in oncological imaging of the University Zurich, and by the Swiss cancer research foundation under grant KFS-5645-08-2022.

### Author Declarations

Ethical approval was obtained from the ethics committee of the University Hospital Zurich on 2025-01-21 and registered under the Swiss Business Administration System for Ethics Committees (BASEC) number 2024-01936. The study will be performed in accordance with the Declaration of Helsinki, the principles of Good Clinical Practice, the Human Research Act (HRA) and the HumanResearch Ordinance (HRO), as well as other relevant local regulations. Written informed consent is obtained from each participant prior to their inclusion in the study.

